# An Explainable Artificial Intelligence based Prospective Framework for COVID-19 Risk Prediction

**DOI:** 10.1101/2021.03.02.21252269

**Authors:** Vishal Sharma, Piyush, Samarth Chhatwal, Bipin Singh

## Abstract

Given the spread of COVID-19 to vast geographical regions and populations, it is not feasible to undergo or recommend the RT-PCR based tests to all individuals with flu-like symptoms. The reach of RT-PCR based testing is still limited due to the high cost of the test and huge population in few countries. Thus, alternative methods for COVID-19 infection risk prediction can be useful. We built an explainable artificial intelligence (AI) based integrated web-based prospective framework for COVID-19 risk prediction. We employed a two-step procedure for the non-clinical prediction of COVID19 infection risk. In the first step we assess the initial risk of COVID19 infection based on carefully selected parameters associated with COVID-19 positive symptoms from recent research. Generally, X-ray scans are cheaper and easily available in most government and private health centres. Therefore, based on the outcome of the computed initial risk in first step, we further provide an optional prediction using the chest X-ray scans in the second step of our proposed AI based prospective framework. Since there is a bottleneck to undergo an expensive RT-PCR based confirmatory test in economically backward nations, this is a crucial part of our explainable AI based prospective framework. The initial risk assessment outcome is analysed in combination with the advanced deep learning-based analysis of chest X-ray scans to provide an accurate prediction of COVID-19 infection risk. This prospective web-based AI framework can be employed in limited resource settings after clinical validation in future. The cost and time associated with the adoption of this prospective AI based prospective framework will be minimal and hence it will be beneficial to majority of the population living in low-income settings such as small towns and rural areas that have limited access to advanced healthcare facilities.

## 1. Introduction

The COVID-19 has affected hundreds of nations around the globe. Countries such as the USA, India, Brazil, and Russia are badly affected given their high populations. There is a huge challenge to perform RT-PCR based confirmatory test for the diagnosis of COVID-19 for a major fraction of people in these countries. Furthermore, issues related to false positives, false negatives, turnaround times and technical difficulty of performing RT-PCR based tests is also reported. (Surkova et al., 2020). Therefore, alternative, and cost-efficient diagnostic strategies for COVID-19 have huge potential not only for the current pandemic but also for our future preparedness to tackle similar situations. We propose an artificial intelligence (AI) based end-to-end web-based prospective framework for COVID-19 infection risk prediction (Figures 1 and 2) that can be considered as a prospective framework for the AI based COVID-19 risk prediction in future. In the first part, we assess the initial risk of infection by carefully analysing the factors related to COVID-19 positive symptoms such as age, gender, BMI, fever, persistent cough, shortness of breath, fatigue, diarrhoea, loss of taste and smell, chest pain, abdominal pain, hoarse voice and delirium. (Carfì et al., 2020) (Zhang et al., 2020) (Dixon et al., 2020) (O’Keefe et al., 2020). Based on the computed risk probability, a further confirmatory prediction can be made based on the analysis of chest X-ray scans, it is implemented in the second part of our proposed prospective framework. We also provided explanation of the prediction for both the parts for our proposed predictive AI based prospective framework.

**Figure 1.**
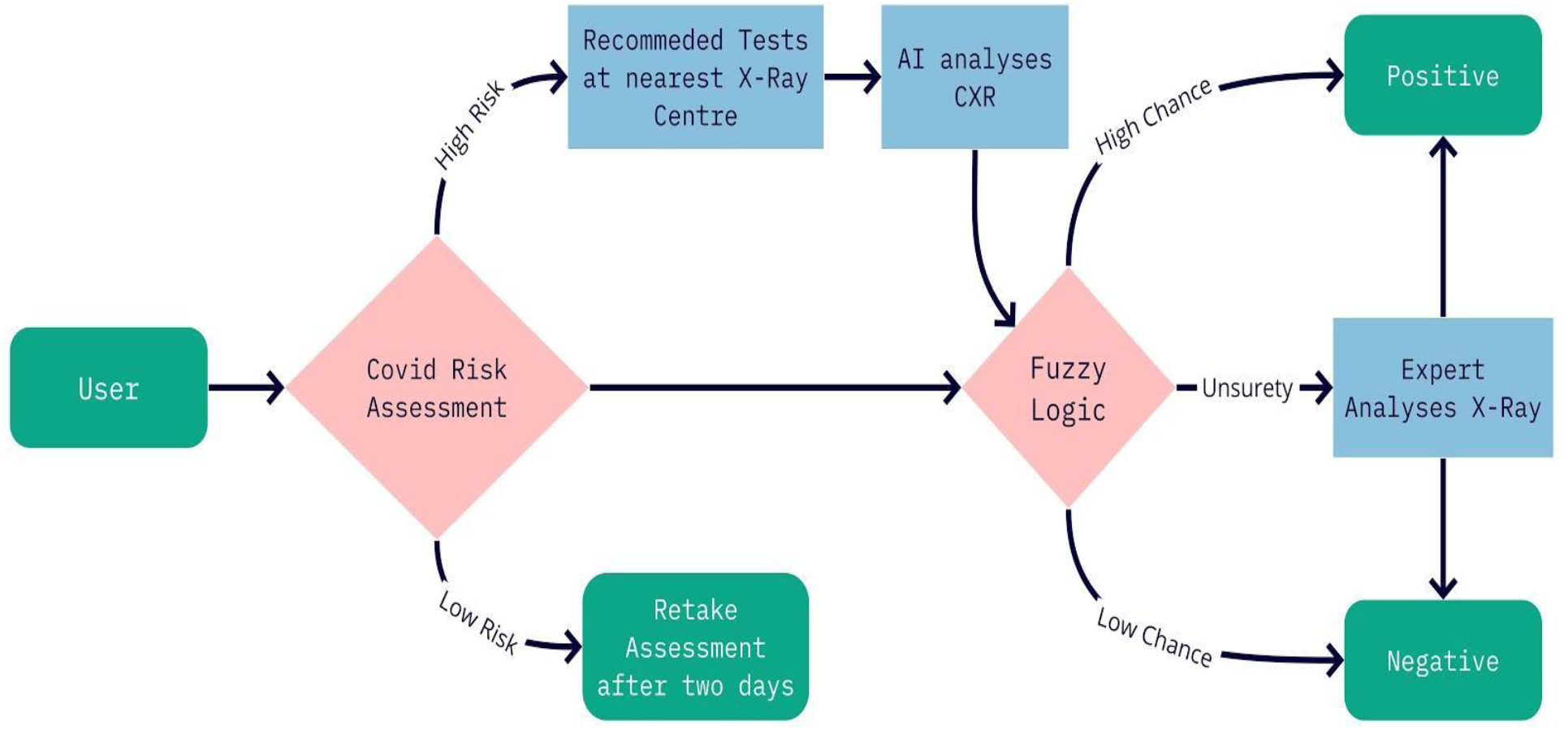
An overview of the integrated web based explainable AI based prospective framework for COVID-19.

**Figure 2.**
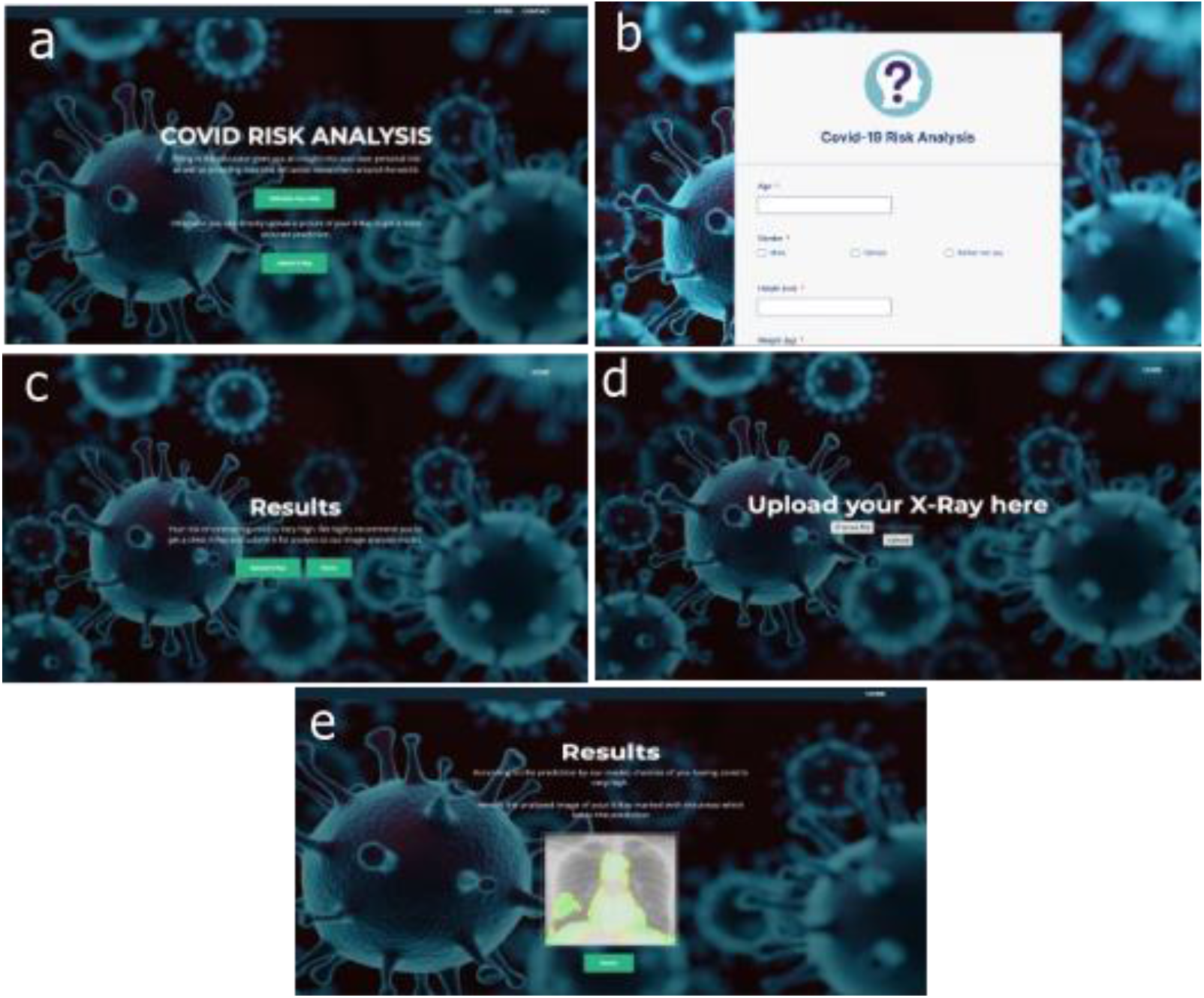
An overview of the different pages of the integrated web application for COVID-19 prediction. a. Home page. b. Risk analysis form. c. Results page for risk analysis form. d. chest X-ray scan image upload. e. chest X-ray prediction result.

Our prospective framework will allow the user to take an online risk assessment test through a web application which will compute the probability of a user to be COVID-19 positive or not based on symptoms suggested for COVID-19, in addition to medical history and recent travel history of the user. Based on the result of this preliminary assessment, a chest X-ray scan is recommended to the user for further confirmation of COVID-19 infection. Once the user takes the chest X-Ray scan, it can be uploaded to the web-based predictive framework by the user, hospital, or the radiology centres in future. The chest X-Ray scans are then analysed by different state-of-the-art deep learning models. The prediction results from the chest X-ray scan and preliminary risk assessment results are passed through the fuzzy predictor which provides the probability of the user to be COVID-19 positive or negative and the final results is shown to the user. The fuzzy predictor will reduce false positives and false negatives of the prediction. The cases where the result seem inconclusive, can be referred to the medical expert for further assessment.

Good medical and diagnostic lab facilities are very limited in developing and low-income countries across the world. The ratio of number of patients and doctor is ever increasing, and the number of quality doctors is decreasing in villages and small towns. Adding the unintentional biases of human nature into the diagnosis and treatment, the end results can be far from ideal. (i.e., variation in diagnosis amongst medical experts). Through this work, we attempt to provide a prospective framework to bridge the gap by automating some of the processes involved in the COVID-19 infection prediction to take some burden off from the shoulders of the healthcare professionals in future.

The first part of this work i.e., assessment of initial risk of COVID-19 infection is based on machine learning for regression and involves hyperparameter tuning. We adopted the best practices for the model selection, algorithm selection and evaluation criteria. (Raschka, 2018). Since conventional methods such as the holdout method for model evaluation and selection are not recommended for small datasets, different types of bootstrap techniques can be used for estimating the uncertainty of performance estimates. Alternative methods, such as the combined F-test 5×2 cross-validation and nested cross-validation, are recommended for comparing machine learning algorithms when datasets are small. We have optimized the hyperparameters associated with each of the models to get the model with optimal predictive ability. (Wu J et al., 2019). Several experiments were conducted on standard test datasets for this purpose.

Since the second part of this work is based on confirmation of COVID-19 infections based on chest X-ray scans, it is important to mention that there are vast number of research papers related to the detection of different chest diseases based on chest X-ray scans. However, most of these papers are based on the NIH Chest X-Ray dataset that has over 100,000 X-Ray scan images of 30,000 different people. The popular artificial neural network based CheXNet model was reported to detect pneumonia at a radiologist level accuracy. (Rajpurkar et al., 2017). It is a 121 layers deep neural network and trained on NIH Chest X-Ray dataset. It can achieve better F1-Score than expert radiologists. This previous work provided the motivation for the present work to integrate an explainable AI based prospective framework for COVID-19 using the chest X-ray scans.

## 2. Methods and Materials

### 2.1 Dataset and Preprocessing

For predicting the risk of COVID-19 infection, we built a machine learning model that was trained on data published by Nexoid (https://www.covid19survivalcalculator.com/en/research). The data is licenced under “Attribution 4.0 International (CC BY 4.0)” license; hence, we could use it for this work. (Nexoid, n.d.). To ensure the anonymity within the dataset, Nexoid has taken the following steps:

- No email addresses are published.
- No IP addresses are published.
- No date of birth year is published.
- No month of birth is published.
- Age is reduced to 10-year bandings.
- Timestamp randomly adjusted (−24 hours to +24 hours)
- IP location longitude and latitude randomly adjusted (−0.1 to +0.1)

In total they have collected 9,12,768 records from their COVID-19 risk calculator (https://www.covid19survivalcalculator.com). Based on their work, they built an algorithm that calculates a score for predicting the risk of COVID-19 infection. However, the algorithm used by Nexoid is not available publicly, hence it is not yet open sourced. Nexoid’s calculator estimates the COVID-19 risk of infection based on various factors such as COVID-19 symptoms, being in nursing home, contact with COVID-19 patient, house person count, working in healthcare services, contact person count in a week, presence of heart disease, kidney disease, liver disease, diabetes, lung disease, travel to school or work and usage of public transport. They stated that factors such as COVID-19 symptoms, nursing home visit, COVID-19 contact, family size, working in healthcare and using public transport are most often positively correlated with COVID-19 infection risk. However, underlying method and model used for this prediction is currently not available.

The objective of our work is to use the important factors for COIVD-19 to accurately calculate the infection risk and simplify the process by excluding correlated (height/weight) and some of the complex factors (government action, government control, government spending, prescription medication, latitude, longitude, survey date and country) from the original dataset. Furthermore, some of these factors are not easy to acquire and patients may not be comfortable in sharing this additional information. Therefore, we have excluded these factors from building the model for COVID-19 risk prediction.

### 2.2 Machine Learning Pipeline for Risk Prediction

We performed exploratory data analysis to identify most important features and correlation between different features. Further we carried out the data preprocessing, such as data cleaning by removing outliers, null values and encoding categorical data for regression models. We split the data into train and validation sets in the ratio of 0.3:0.7. We selected following six machine learning models for training:

1. Linear Regression
2. Decision Tree
3. Random Forest
4. KNN
5. XGBoost
6. AdaBoost

Since we have features with values at different scales, it is important to standardize them to the same range so that the algorithm does not weigh greater values more and smaller values less, regardless of the unit of the values. This is crucial for distance-based algorithms such as KNN, SVM, linear regression, etc. We choose RobustScaler (Cao et al., 2016) for scaling, since it is based on quantile range and therefore not easily influenced by outliers.

For optimizing linear regression models, we removed the features which failed to explain mortality risk which is our target variable. For this purpose, we used a backward elimination method and deselected features that have p-value more than 0.05. For optimizing other machine learning models, we used grid search cross validation for searching hyperparameters and selecting the best hyperparameters. After training the models on best hyperparameters, we evaluated the models using four different metrics. (Emmert-Streib et al., 2019).

1. R^2^.
2. Adjusted R^2^.
3. Root Mean Square Error.
4. Calculating statistical significance between models using 5×2 CV using MLxtend package. (Raschka, 2018).

Based on the above metrics we selected XGBoost Model. (Chen et al., 2016).

### 2.3 Deep learning based chest X-ray analysis

For Chest X-xay based prediction, we collected COVID-19 X-ray images from open-source data collected by Cohen et al. (Cohen et al., 2020). We could only find 184 COVID-19 X-ray images (when this work was performed) and around 700 X-Ray images for other pneumonia infection. We collected more non-COVID images from CheXpert Dataset. (Irvin et al., 2019). We had 3000 non-COVID images and 100 COVID-19 images in the train dataset. In the test dataset we had 580 non-COVID images and 84 COVID-19 images. Since the training dataset was highly imbalanced, we had to use more oversampling techniques to increase COVID-19 images (Figure 3). We used image augmentation techniques such as flip, mirror, unsharp and gaussian blur filter (fif, M., Said, Y., & Atri, M. (2020). After oversampling COVID-19 images, we had 1600 COVID-19 X-Ray images and 3000 non-COVID images.

**Figure 3:**
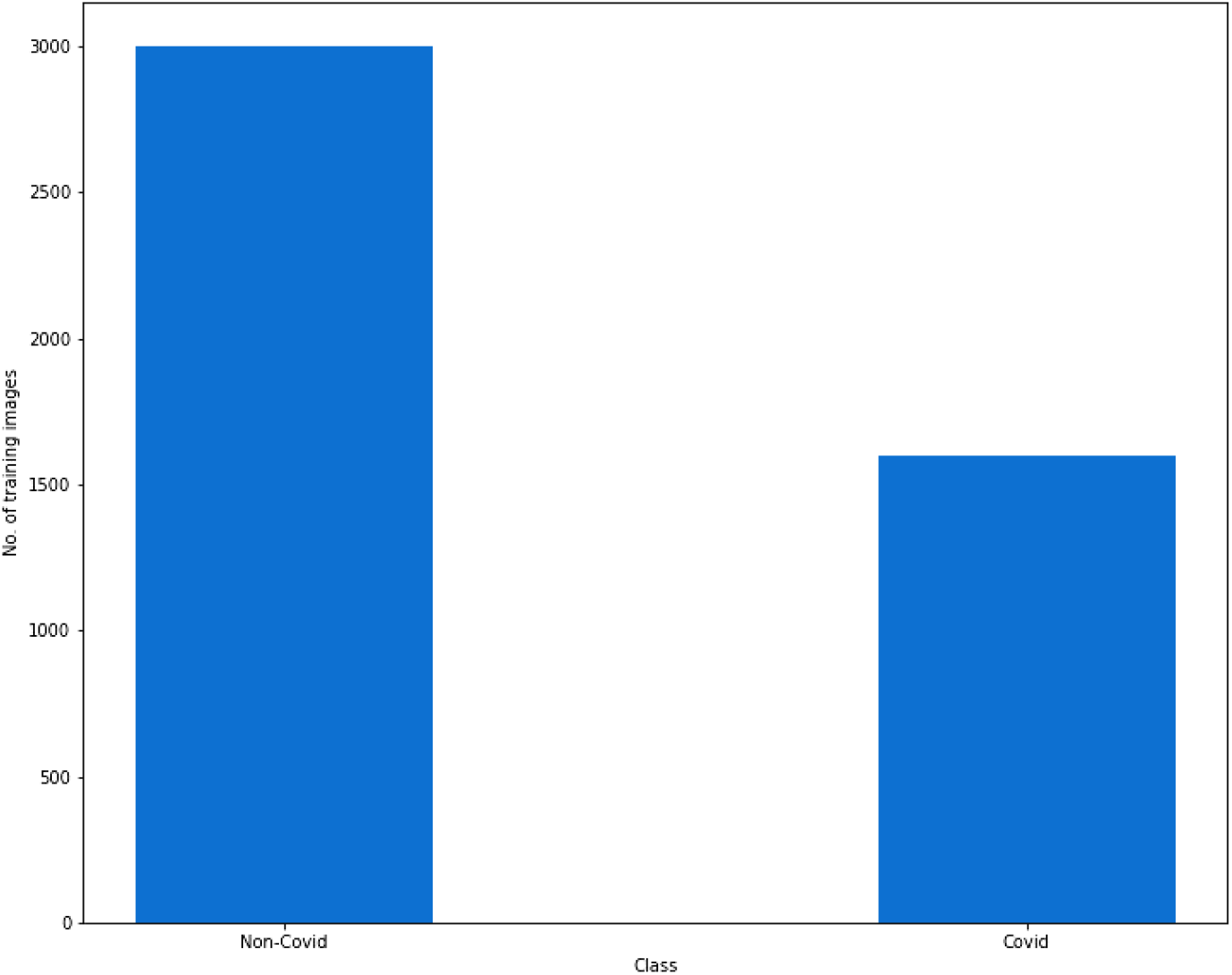
Number of training images after oversampling

We built and trained 3 different models to compare the predictive performance. First, we trained InceptionV3 model (Figure 4; Szegedy et al., 2016) using transfer learning technique. We tried training the Inception model with various configurations like training the whole network from scratch, training only fully connected layers, training the last few layers, etc. The best result was in the case where we initialized the model with InceptionV3 ImageNet weights and set all the layers as trainable. We used SoftMax as activation function and categorical cross entropy loss as loss function with Adam optimizer.

**Figure 4.**
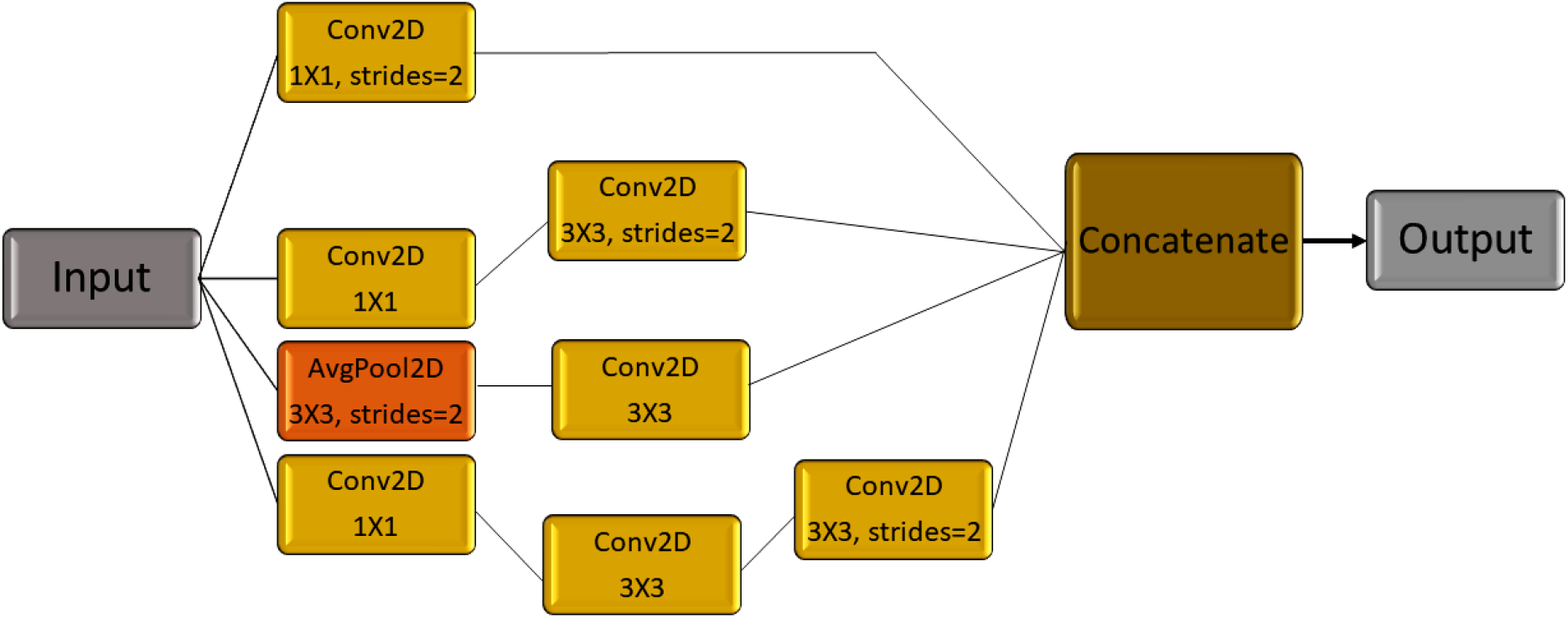
An overview of InceptionV3 model architecture.

Next, we tried the VGG16 model (Figure 5; Simonyan et al., 2014) using transfer learning techniques. Similarly, we tried training the VGG16 model with various configurations like training the whole network from scratch, training only fully connected layers, training the last few layers, etc. The best result was in the case where we initialized the model with VGG16 ImageNet weights and set all the layers as trainable. We used SoftMax as activation function and categorical cross entropy loss as loss function with Adam optimizer.

**Figure 5.**
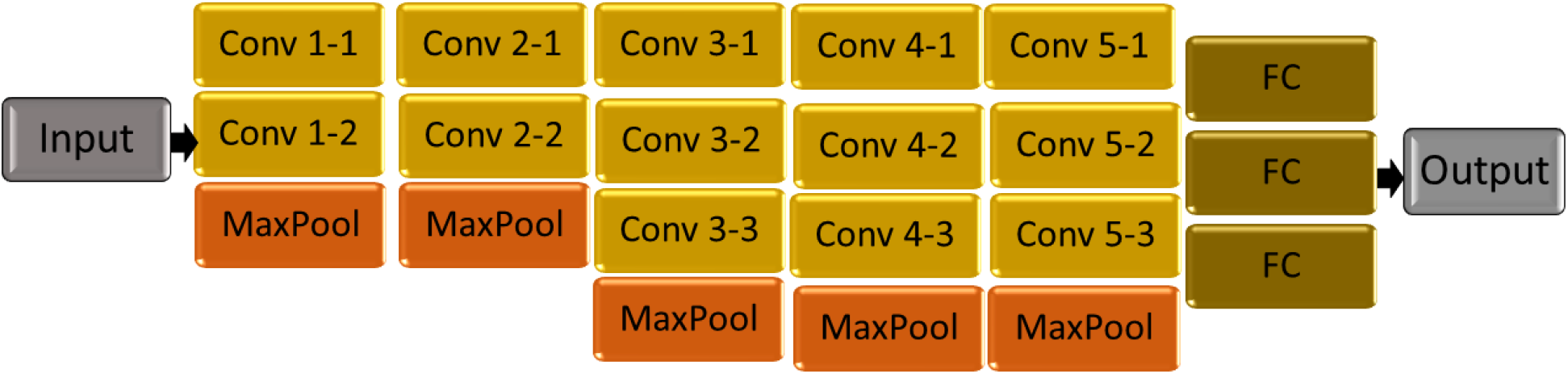
An overview of the VGG16 model architecture.

### 2.4 Custom 3-layer CNN model

Next, we built a custom CNN model (Figure 6) with dropout and SoftMax activation function at the output. It has three convolution layers with rectified linear unit (ReLU) activation function. We used Adam optimizer to train the model.

**Figure 6.**
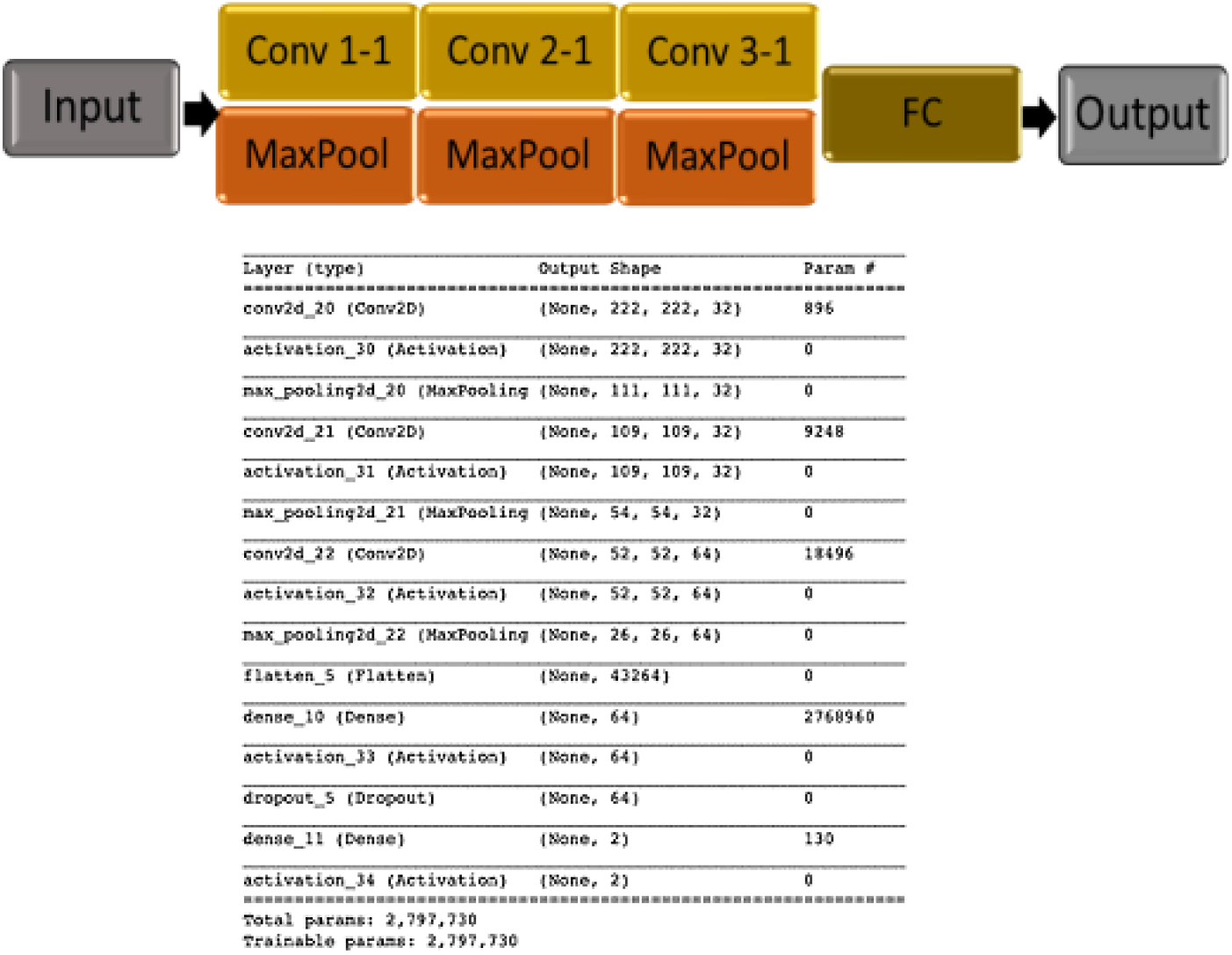
Custom CNN model.

We also explained the model predictions on chest X-ray scans by using LIME (Local Interpretable Model-Agnostic Explanations). (Ribeiro et al., 2016). We used LIME to provide explanations for the prediction made by the deep neural networks on X-ray scans. LIME algorithm can explain the prediction with confidence by approximating the deep neural network models locally with an interpretable model. Thus, the reason behind any of the prediction made is available to the user of our proposed model framework that is highly important for a healthcare professional to take an informed decision.

We evaluated all three models on a test dataset of 580 non-COVID images and 84 COVID-19 images. We compared the models based on precision, recall and accuracy for both the classes.

### 2.5 Web Application development

The web application has been built using the Django framework and Python programming language to deploy the machine learning model for taking inputs and return the prediction results. Django framework provides the necessary layout for a web application that can be changed according to the requirement. The frontend part was written in HTML5, CSS and JavaScript. The frontend and the backend part were combined to provide an integrated and simple interface to the users (Figure 7). The two parts in the backend, namely, COVID-19 risk analysis part, which deals with finding out the chance of a user contracting COVID-19 based on his routine and other parameters. The second part is where a user or healthcare professional can upload the chest X-ray scan to get a confirmatory prediction of advanced stage COVID-19 infection for further clinical intervention.

**Figure 7.**
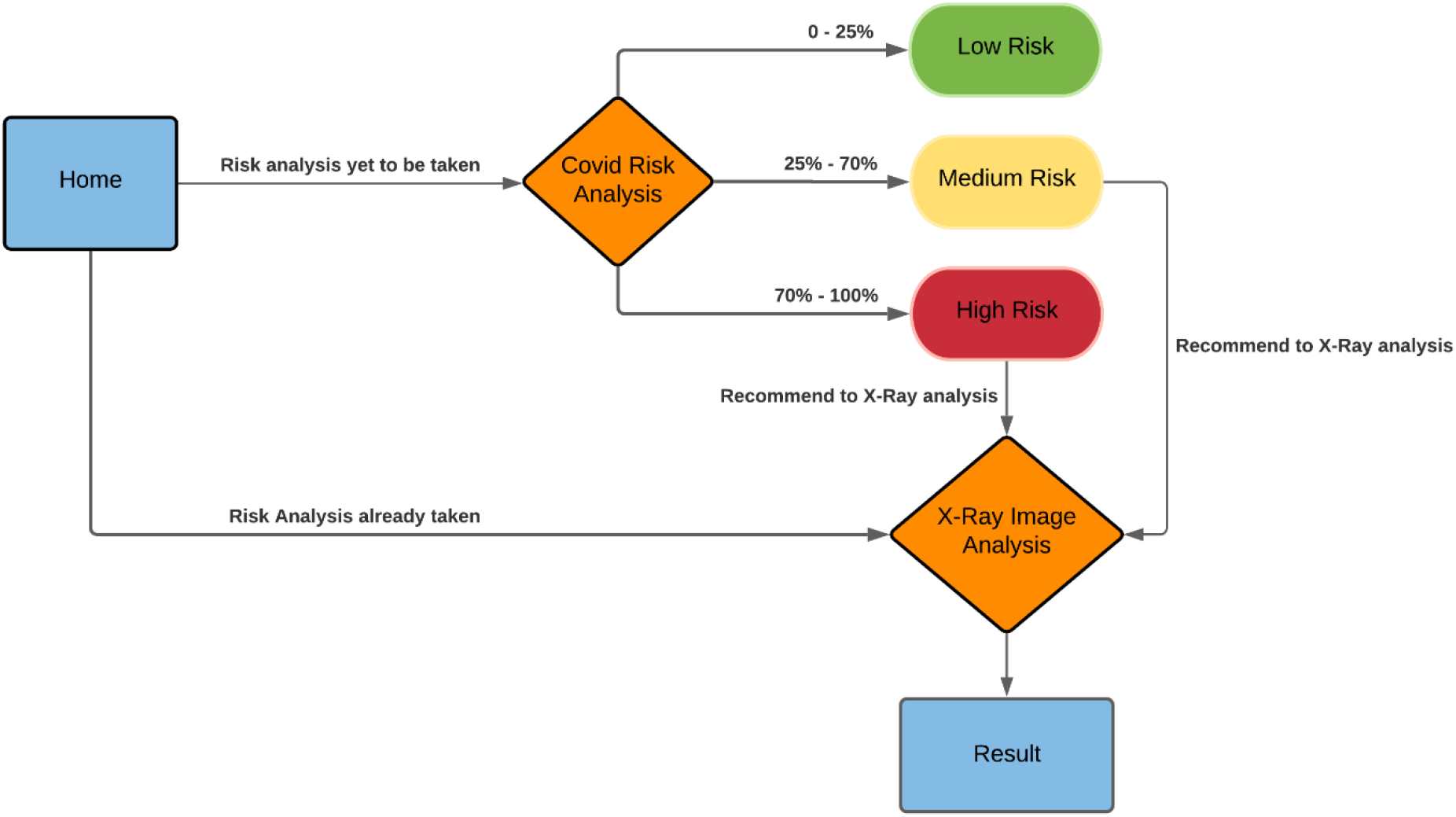
An overview of the developed integrated web-based model framework for COVID-19 risk prediction.

## 3. Results and Discussion

### 3.1 Performance of different models for COVID-19 risk assessment

Based on performance and accuracy (Figures 8-9) of different models, we found that the XGboost and Random forest based regression models were the best and therefore can be used for making accurate prediction about COVID-19 infection risk. Further we analysed the explainability of the Random Forest regression model using LIME and found that the factors such as COVID-19 symptoms, COVID-19 contact, nursing home visit, working in healthcare, public transport usage, work travel critical, heart disease, work travel non-critical, liver disease and age were the factors used for making the predictions.

**Figure 8.**
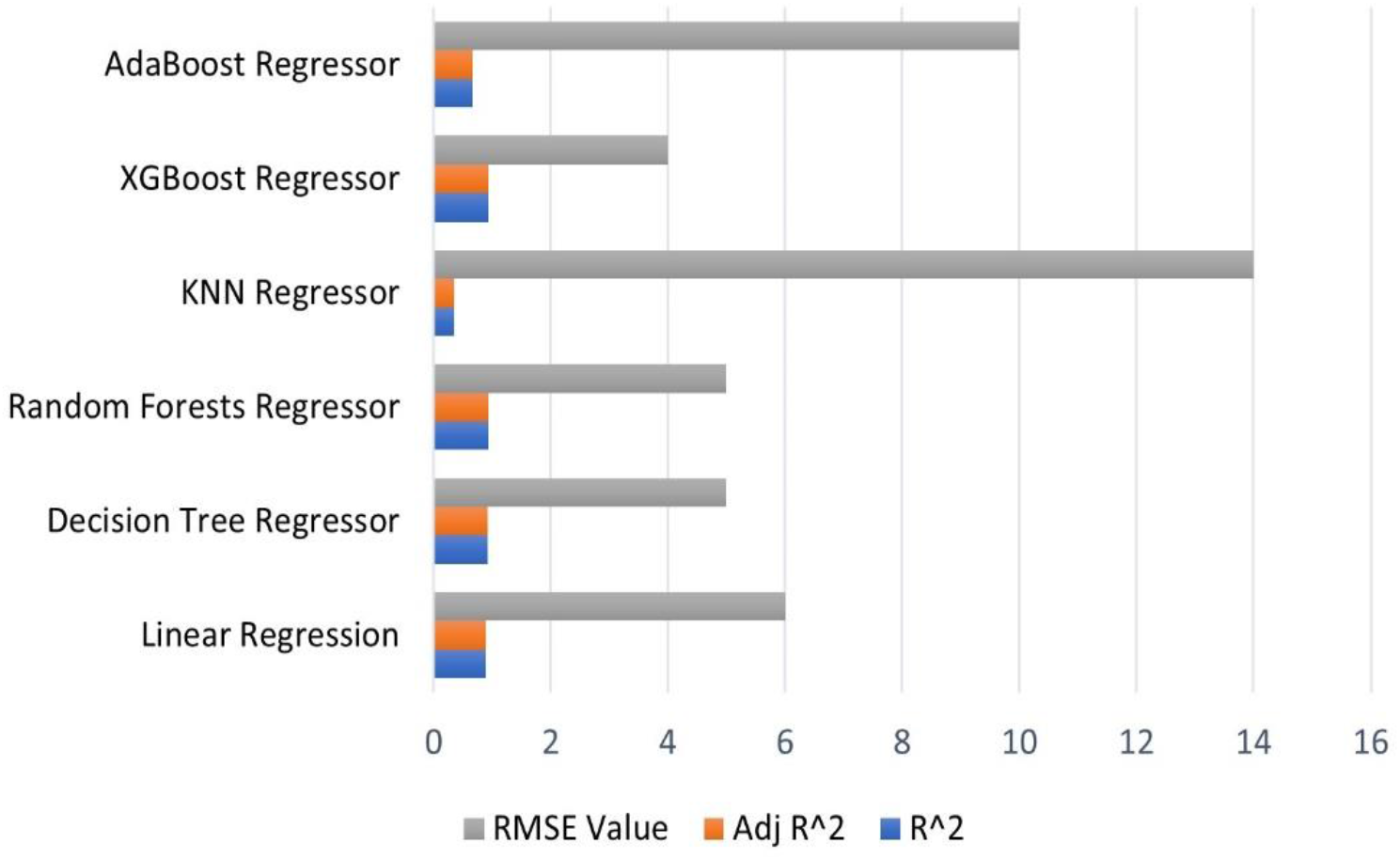
Comparison of performance of different methods used for building the machine learning model.

**Figure 9.**
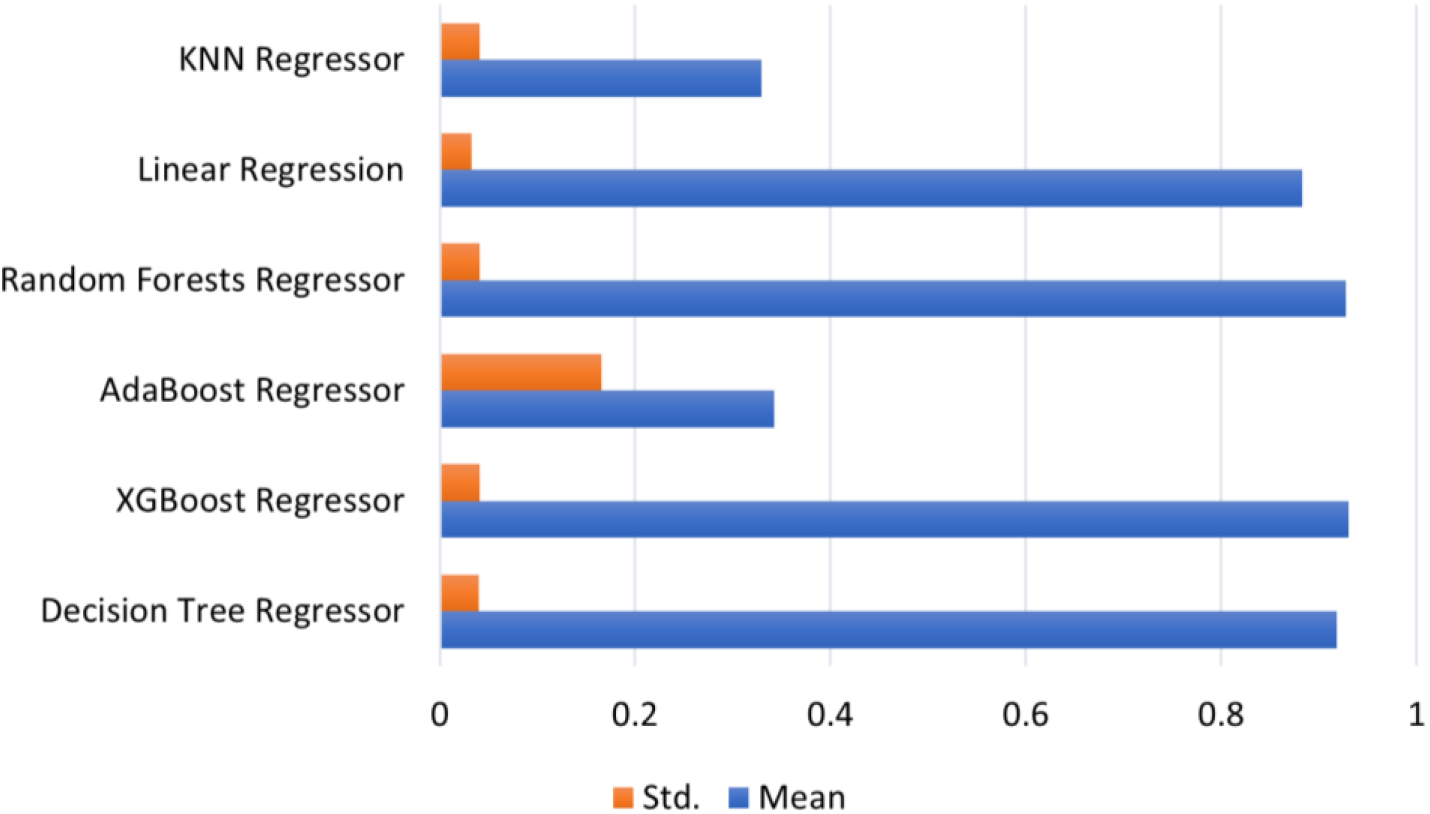
Comparison of the accuracy of different machine learning models based on R^2 value across each cross-validation fold.

### 3.2 Performance of different models on chest X-ray scans for COVID-19 prediction

The performance of the Inception V3 and VGG 16 deep learning models were comparable with the custom three-layer CNN model (Figure 10 and Table 1). Thus, it is notable that a relatively very simple three-layer CNN model could classify COVID-19 positive and negative chest X-ray scans with precision similar to state-of-the-art deep learning models. This suggest that in case of smaller dataset, a simple CNN model can be effectively used for complex classification tasks if trained appropriately. We further provided the LIME explanation for the prediction made by the custom CNN model to further assess and explain the quality of the prediction. It was also found that the LIME mostly concentrated on the pixels of the lung region to provide explanation for the classification of the chest X-ray scans as COVID-19 positive or COVID-19 negative (Figure 11).

**Table 1.** Performance of different deep learning models of test set chest X-ray scans (C:COVID-19 positive and NC: COVID-19 negative)

|  | Inception V3 |  | VGG 16 |  | Custom CNN |  |
| --- | --- | --- | --- | --- | --- | --- |
| **Accuracy** | 0.98 |  | 0.99 |  | 0.99 |  |
| **Precision** | 0.94 (C) | 0.98 (NC) | 0.99 (C) | 0.99 (NC) | 0.94 (C) | 1.0 (NC) |
| **Recall** | 0.88 (C) | 0.99 (NC) | 0.94 (C) | 1.0 (NC) | 0.99 (C) | 0.99 (NC) |
| **F1-score** | 0.91 (C) | 0.99 (NC) | 0.96 (C) | 0.99 (NC) | 0.97 (C) | 0.99 (NC) |

**Figure 10.**
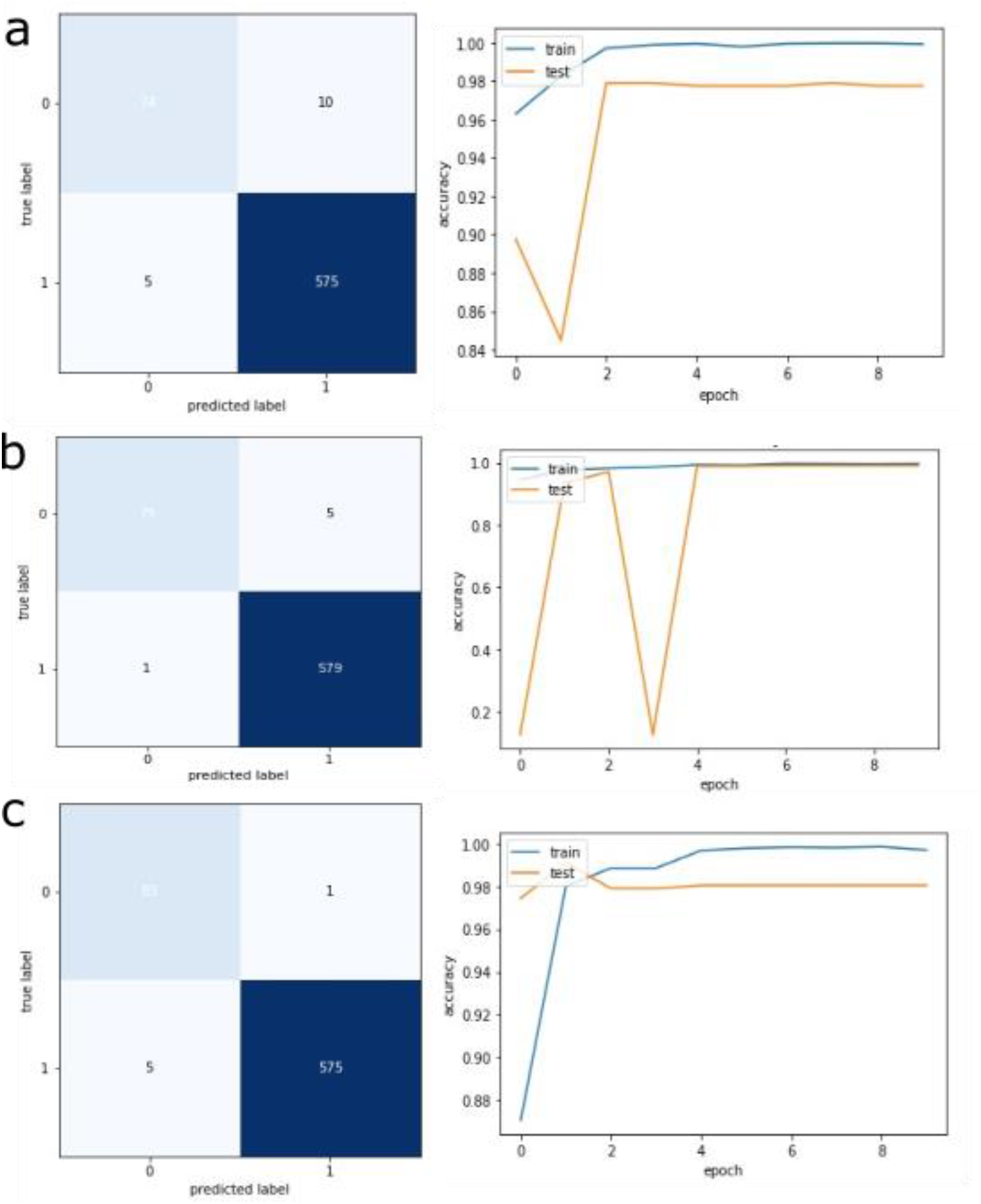
Confusion matrices and performance curves on training and test datasets for different deep learning models. a. InceptionV3 model b. VGG16 model and c. Custom CNN model. The horizontal axis of the curves shows the number of epochs, and the vertical axis shows the accuracy.

**Figure 11.**
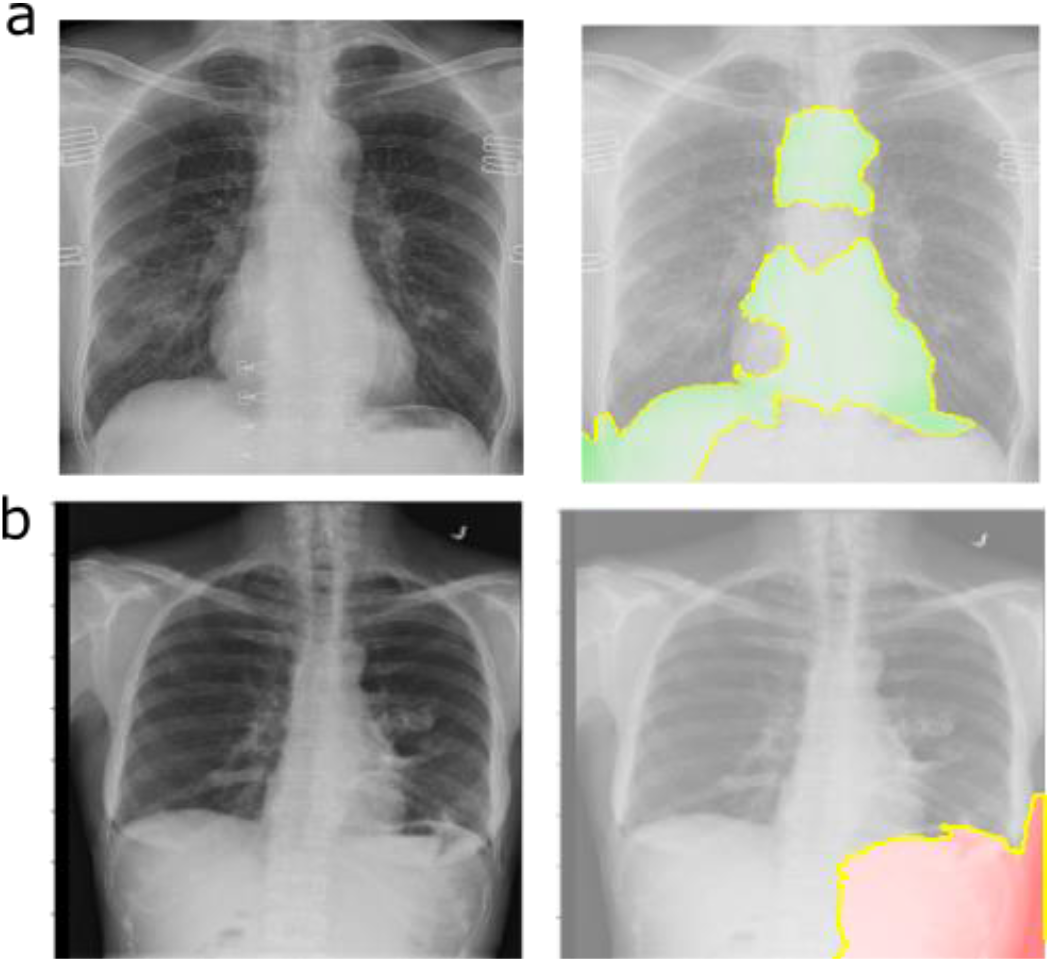
LIME explanation for the prediction made by custom CNN model on (a) COVID-19 positive and (b) COVID-19 negative chest X-ray scans.

## 4. Conclusions

We developed a web-based prospective framework for non-clinical prediction of COVID-19 infection risk and further assessment of infection using chest X-ray scans. This integrated web-based framework serves an example that can be adopted in future for building similar clinical framework to assess the risk of COVID-19 infection and diagnosis in future after clinical validation. The utility of such framework will be primarily for the population living in remote locations where it is sometimes difficult to take RT-PCR based confirmatory test either due to its availability or cost. Furthermore, such framework can also be used for purely research purpose or for a non-clinical assessment of COVID-19 infection risk using important factors provided by the user and chest X-ray scan. This prospective framework can be further enhanced and improved for its utility to enable it to reach the stage of clinical usage in future after clinical validation. The developed web-based application can be easily hosted on a public domain for the purpose of further research and improvement.

## Data Availability

All the data used in this work is available in the public domain.

## Authors’ contributions

VS, Piyush, SC and BS: The conception and design of the study, or acquisition of data, or analysis and interpretation of data.

VS, Piyush, BS: Drafting the article or revising it critically for important intellectual content. VS, Piyush, SC and BS: Final approval of the version to be submitted.

## Statement on conflicts of interest

There is no conflict of interest to declare.

## Notes

### Competing Interest Statement

The authors have declared no competing interest.

### Funding Statement

No funding is received for this work.

